# Associations of Adipokine Levels with Levels of Remnant Cholesterol: the Multi-Ethnic Study of Atherosclerosis (MESA)

**DOI:** 10.1101/2023.04.24.23289072

**Authors:** Renato Quispe, Ty Sweeney, Seth S. Martin, Steven R. Jones, Matthew A. Allison, Matthew J. Budoff, Chiadi E. Ndumele, Mohamed B. Elshazly, Erin D. Michos

## Abstract

**Background:** The metabolic syndrome phenotype of individuals with obesity is characterized by elevated levels of triglyceride (TG)-rich lipoproteins and remnant particles, which have been shown to be significantly atherogenic. Understanding the association between adipokines, endogenous hormones produced by adipose tissue, and remnant cholesterol (RC) would give insight into the link between obesity and atherosclerotic cardiovascular disease.

**Methods:** We studied 1,791 MESA participants of an ancillary study on body composition who had adipokine levels measured (leptin, adiponectin, resistin) at either visit 2 or 3. RC was calculated as non-high density lipoprotein cholesterol minus low-density lipoprotein cholesterol (LDL-C), measured at the same visit as the adipokines, as well as subsequent visits 4 through 6. Multivariable-adjusted linear mixed effects models were used to assess the cross-sectional and longitudinal associations between adipokines and levels of RC.

**Results:** Mean (SD) age was 64.5±9.6 years and for body mass index (BMI) was 29.9±5.0 kg/m2; 52.0% were women. In fully adjusted models that included BMI, LDL-C and lipid-lowering therapy, for each 1-unit increment in adiponectin, there was 14.4% (12.0, 16.8) lower RC. With each 1-unit increment in leptin and resistin, there was 4.5% (2.3, 6.6) and 5.1% (1.2, 9.2) higher RC, respectively. Lower adiponectin and higher leptin were also associated with longitudinal increases in RC levels over median follow-up of 5(4-8) years.

**Conclusions:** Lower adiponectin and higher leptin levels were independently associated with higher levels of RC at baseline and longitudinal RC increase, even after accounting for BMI and LDL-C.

**CLINICAL PERSPECTIVE:** *What is new?:* - Among individuals without history of cardiovascular disease, adiponectin is inversely associated with cross-sectional levels of remnant cholesterol, whereas leptin and resistin are directly associated. - Adiponectin had an inverse association with progression of remnant cholesterol levels over time.

*What are the clinical implications?:* - Adiponectin levels were not associated with LDL-C levels but with levels of triglyceride-rich lipoproteins, particularly remnant cholesterol. -Incrementing adiponectin via lifestyle modification and/or pharmacological therapies (i.e. GLP-1 agonists) could be a mechanism to reduce remnant cholesterol levels and ultimately cardiovascular risk.

## INTRODUCTION

Obesity is one of the most important risk factors for development of atherosclerotic cardiovascular disease (ASCVD)^1-3^ and it is projected that 1 in 2 U.S. adults will be classified as having obesity (defined by a body mass index (BMI) ≥ 30 kg/m^2^) by 2030.^4^ The pathophysiologic link between obesity and ASCVD is not fully understood.^5^ In this regard, the metabolic syndrome lipid phenotype of individuals with obesity is often characterized by elevation of triglyceride (TG) levels and low levels of high density lipoprotein cholesterol (HDL-C).^6^

Mounting evidence from the last decade has shown the role of TG and TG-rich lipoproteins (TGRL) in the development of ASCVD.^7^ Remnant lipoprotein particles (RLPs) are partially lipolysed lipoproteins derived from TGRL of both liver [very low-density lipoprotein, (VLDL)] and intestinal (chylomicron) origins,^8^ which have been shown to contribute to atherosclerosis independent of, and additional to, low-density lipoprotein cholesterol (LDL-C).^9, 10^ These remnant particles contain a large amount of cholesterol and contribute to endothelial dysfunction, inflammation and ultimately atherogenesis.^7, 11^ Interestingly, elevated levels of remnant cholesterol are strongly associated with higher ratios of TG to HDL-C,^12^ which are often encountered among individuals with obesity and metabolic syndrome. The association between remnant cholesterol (RC) and ASCVD has been established by genetic^13, 14^ as well as observational studies.^15, 16^

Adipokines, such as leptin, adiponectin and resistin, are endogenous hormones that are released from adipose tissue or adjacent inflammatory cells and have been shown to influence several metabolic processes such as insulin sensitivity, endothelial function and appetite regulation.^17-21^ However, the association of adipokines with levels of RC has not been fully elucidated, as well as the role of different adipokines in the prediction of change in RC levels. In the analysis described below, we evaluated the associations between endogenous adipokines and 1) cross-sectional levels of RC; and 2) progression of RC levels over time.

## METHODS

### Study population

The Multi-Ethnic Study of Atherosclerosis (MESA) cohort enrolled 6,814 men and women aged 45 to 84 years who were free of CVD at baseline (2000-2002). Adipokine levels and the computed tomography (CT) assessment of visceral and subcutaneous adipose tissue (VAT and SAT) were measured as part of an ancillary study in 1,970 men and women at either visit 2 or visit 3 (randomly assigned). The baseline for this analysis was the time of the adipokine measurement (either visit 2 or 3). We included all MESA participants who had endogenous adipokines (leptin, adiponectin, resistin) and a standard lipid panel obtained at either visit 2 or 3. For the primary cross-sectional analysis, we used the lipid panel that was at the same visit as the adipokine measurements, but for longitudinal analysis, we also used lipid panels from subsequent visits. Individuals with missing data on the exposure variables, the outcome variables, or the covariates in our model were excluded (n=179), leaving a primary analytical sample of 1,791 individuals.

### Independent variables assessment

The primary independent variables were levels of the endogenous adipokines (leptin, adiponectin, and resistin) and their ratios (leptin/adiponectin and resistin/adiponectin). Samples of fasting serum were obtained at either visit 2 (2002-2004) or visit 3 (2004-2005), at the visit of the abdominal CT scan,^22-24^ and immediately frozen at −70°C. In 2009, the adipokines (adiponectin, leptin, and resistin) were measured from these stored serum samples using a Bio-Rad Luminex flow cytometry (Millipore, Billerica, MA) at the Laboratory for Clinical Biochemistry Research (University of Vermont, Burlington, VT), as previously reported.^25, 26^ The coefficients of variation for these assays ranged from 6 to 13%.

### Dependent variables assessment

From a standard lipid panel, RC levels were estimated as non-HDL-C *minus* LDL-C. LDL-C was estimated using the Martin/Hopkins equation. This method estimates LDL-C using 1 of 174 different factors for the TG to VLDL-C ratio according to non-HDL-C and TG levels when TG levels are <400 mg/dL.^27^ We additionally performed similar estimation when TG levels were 400-799 mg/dL using an expanded version of the Martin/Hopkins method that uses several more factors with increased accuracy.^28^ Our group has previously shown that this estimation method for RC levels is more accurate than using the Friedewald equation.^29^

### Covariates

The covariates included in this study include the following demographic, behavioral and ASCVD risk factors measured at the visit of the adipokine assessment: age, sex, race/ethnicity, study site, education (<high school; high school or vocational school; college, graduate or professional school), cigarette smoking status (current, former or never), physical activity (MET hrs/week of moderate or vigorous activity, continuous), BMI (in kg/m^2^), total cholesterol (mg/dL), HDL-C (mg/dL), use of lipid lowering medications (yes/no), diabetes (defined as fasting blood sugar ≥ 126 on non-fasting glucose ≥ 200 mg/dl or medication use), and measures of abdominal body composition (VAT and SAT) by CT. In exploratory models, we replaced BMI with VAT and SAT measures from abdominal CTs from visits 2/3. We also explored new use of lipid lowering medications during follow-up at visits 4, 5 and 6.

### Statistical analysis

We described the baseline demographics and clinical characteristics of the study participants at the time of their adipokine level measurement, stratified by tertiles of each adipokine (adiponectin, leptin and resistin, separately). Continuous variables were expressed as mean and standard deviations (SD) and categorical variables as frequency and percentages. For our regression models, both RC and adipokine levels and their ratios were log-transformed given their non-normal distribution. The regression results were then exponentiated and shown as percent differences using the following formula: (ε^β^-1)*100.

Our primary outcome was RC levels (estimated as described above) assessed cross-sectionally at visits 2 or 3. Our secondary outcome was the prospective change in RC levels from the time of measurement of adipokines (either visit 2 or 3) to Exam 6. We used standard lipid panels from visits 2, 3, 4, 5, and 6 to leverage all available lipid panels in mixed effects linear regression models.

We assessed the longitudinal change in RC levels associated with each of the adipokine levels separately by using multivariable-adjusted linear mixed effects models allowing for random variations in baseline RC levels and longitudinal slope for RC progression across participants. The mixed effects model for longitudinal data leverages all available lipid panel (RC) information from all participants, including those without follow-up measurements, to jointly model the level of RC at baseline and RC change over time. In this mixed effect model, cross-sectional associations are represented by coefficients of the adipokines, which estimate the difference in RC at baseline by varying adipokine levels. Longitudinal associations are represented by coefficients of interactions between adipokines and time since baseline, which estimate the rate of change in RC levels associated with adipokines.

The linear regression models were progressively adjusted as follows: Model 1 – age, sex, race/ethnicity, and study site; Model 2 – Model 1 + education, smoking status, and physical activity; Model 3 – Model 2 + BMI; Model 4 – Model 3 + LDL-C; Model 5 – Model 4 + use of lipid-lowering therapy; Model 6 – Model 5 + diabetes mellitus; Model 7 - Model 5 + subcutaneous fat area, in cm2 (instead of BMI); Model 8 – Model 5 + visceral fat area, in cm2 (instead of BMI) For sensitivity analyses, we evaluated interactions by sex, diabetes mellitus and obesity status (BMI ≥30 vs <30 kg/m2).

## RESULTS

### Study population

The mean age (SD) of the population was 64.5 (9.6) years, and about half of them were women (52%). In terms of race/ethnicity, the majority of individuals were of White (39.8%), followed by Hispanic (25.5%), Black (21.1%) and Chinese-Americans (13.6%). The mean (SD) BMI of the population was 29.9 (5.0) kg/m2. The median (IQR) TG levels were 113 (77-161) mg/dL, and the median (IQR) RC levels were 21.8 (16.9-27.8) mg/dL. The median (IQR) values for adipokines were: adiponectin, 17.5 (11.9-26.4) ng/mL; leptin, 13.1 (5.6-28.2) ng/mL; and resistin, 14.9 (11.9-18.8) ng/mL.

### Baseline characteristics by adipokine levels

The baseline characteristics were estimated per tertiles of each adipokines separately to more evenly distribute the population, and are shown in **Table 1** (adiponectin), **Table 2** (leptin), and **Table 3** (resistin).

**Table 1.** – Baseline characteristics by tertiles of adiponectin (n=1,791)

|  | Lowest Tertile | Second Tertile | Highest Tertile | p-value |
| --- | --- | --- | --- | --- |
| **Age, years** | 62.2 ± 9.2 | 64.5 ± 9.5 | 66.8 ± 9.5 | 0.591 |
| **Female** | **210 (35.2)** | **300 (50.3)** | **421 (70.5)** | **<0.001** |
| **Race/ethnicity** |  |  |  | **<0.001** |
| White | **158 (26.5)** | **225 (37.7)** | **330 (55.3)** |  |
| Chinese-American | **119 (19.9)** | **71 (11.9)** | **53 (8.9)** |  |
| Black | **171 (28.6)** | **115 (19.3)** | **92 (15.4)** |  |
| Hispanic | **149 (25.0)** | **186 (31.2)** | **122 (20.4)** |  |
| **Smoking status** |  |  |  | **0.023** |
| Never | **265 (44.4)** | **272 (45.6)** | **302 (50.6)** |  |
| Former | **246 (41.2)** | **258 (43.2)** | **242 (40.5)** |  |
| Current | **86 (14.4)** | **67 (11.2)** | **53 (8.9)** |  |
| **BMI, kg/m2** | 29.0 ± 5.0 | 28.2 ± 4.9 | 26.4 ± 4.8 | 0.523 |
| **SBP, mmHg** | 124.2 ± 20.5 | 123.6 ± 19.9 | 123.3 ± 22.1 | **0.031** |
| **Triglycerides, mg/dl** | **130 (93-180)** | **117 (81-164)** | **89 (65-136)** | **<0.001** |
| **RC, mg/dl** | **23.6 (19.1-30.2)** | **22.3 (17.3-28.8)** | **18.8 (15-24.9)** | **<0.001** |
| **Total Cholesterol, mg/dl** | 185.6 ± 36.0 | 191.0 ± 34.6 | 193.5 ± 35.4 | 0.632 |
| **LDL-C, mg/dl** | 115.1 ± 31.1 | 115.5 ± 30.2 | 112.5 ± 30.7 | 0.764 |
| **HDL-C, mg/dL** | **44.8 ± 11.2** | **51.3 ± 13.2** | **60.0 ± 16.8** | **<0.001** |
| **Diabetes** | **112 (18.8)** | **72 (12.1)** | **54 (9.1)** | **<0.001** |
| **Antihypertensive medications** | 251 (43.3) | 244 (42) | 246 (42.3) | 0.901 |
| **Lipid-lowering medications** | 147 (25.3) | 152 (26.2) | 133 (22.9) | 0.406 |

Values are shown using mean ±SD for normal and median (25<sup>th</sup>-75<sup>th</sup> percentile) for not-normal continuous variables, and n(%) for categorical variables.
Comparison was made using chi-square (categorical), t-test (normal) or Kruskal-Wallis (not-normal). **Bold** values are statistically significant (p<0.05)
BMI: body mass index, SBP: systolic blood pressure, RC: remnant cholesterol; LDL-C: low-density lipoprotein cholesterol, HDL-C: high-density lipoprotein cholesterol

**Table 2.**
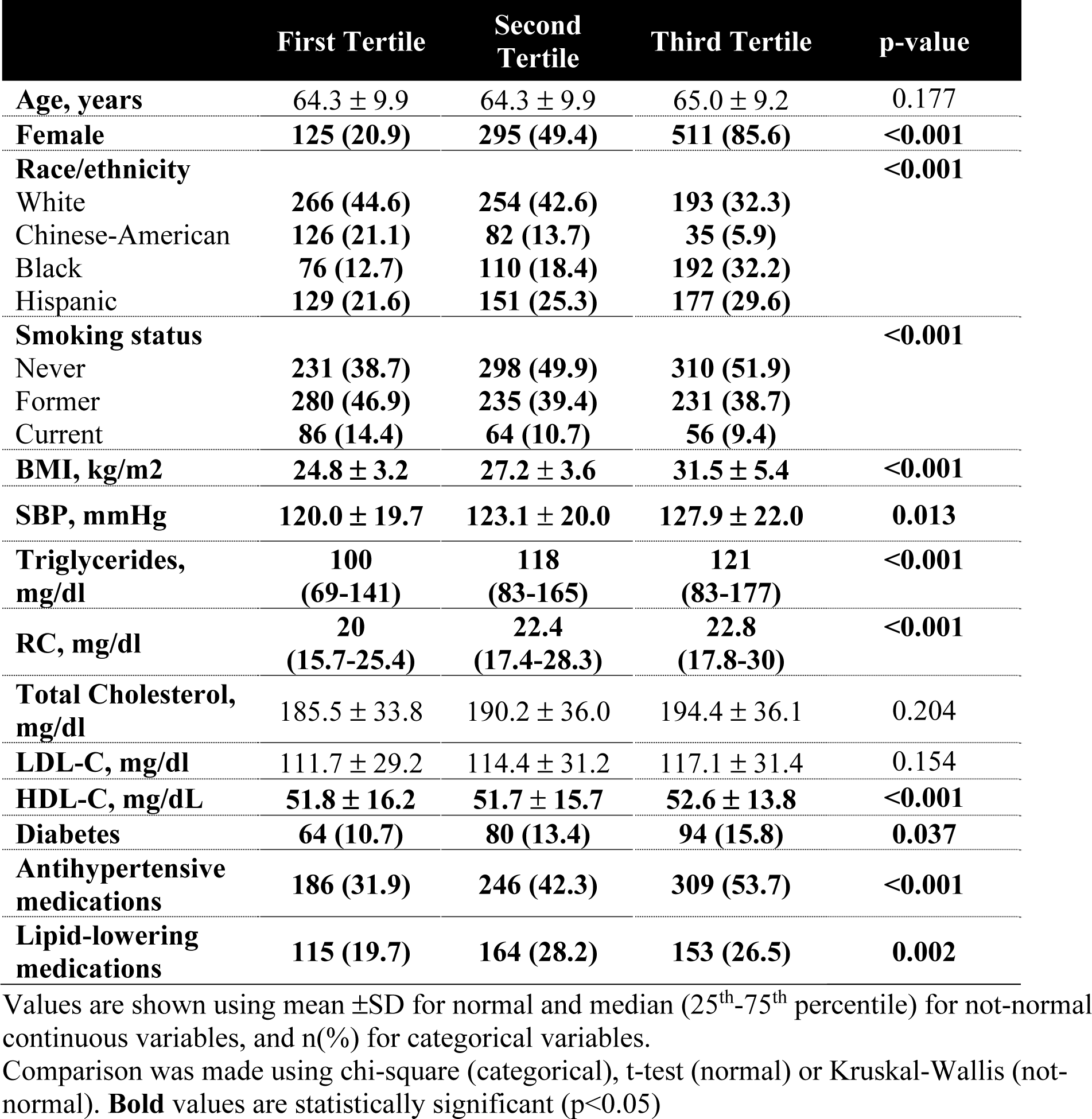
– Baseline characteristics by tertiles of leptin (n=1,791)

**Table 3.** – Baseline characteristics by tertiles of resistin (n=1,791)

|  | First Tertile | Second Tertile | Third Tertile | p-value |
| --- | --- | --- | --- | --- |
| Age, years | 62.8 ± 9.0 | 64.0 ± 9.5 | 66.8 ± 9.7 | 0.139 |
| Female | 307 (51.4) | 292 (48.9) | 332 (55.6) | 0.065 |
| Race/ethnicity |  |  |  | <0.001 |
| White | **209 (35.0)** | **278 (46.6)** | **226 (37.9)** |  |
| Chinese-American | **105 (17.6)** | **64 (10.7)** | **74 (12.4)** |  |
| Black | **122 (20.4)** | **103 (17.3)** | **153 (25.6)** |  |
| Hispanic | **161 (27.0)** | **152 (25.5)** | **144 (24.1)** |  |
| Smoking status |  |  |  | 0.068 |
| Never | 298 (49.9) | 263 (44.1) | 278 (46.6) |  |
| Former | 242 (40.5) | 249 (41.7) | 255 (42.7) |  |
| Current | 57 (9.6) | 85 (14.2) | 64 (10.7) |  |
| BMI, kg/m2 | **27.1 ± 4.7** | **28.1 ± 4.8** | **28.4 ± 5.4** | **0.002** |
| SBP, mmHg | 122.3 ± 20.4 | 122.1 ± 20.5 | 126.6 ± 21.3 | 0.550 |
| Triglycerides, mg/dl | 109<br>(76-149) | 113<br>(76-164) | 117<br>(80-171) | **0.0371** |
| RC, mg/dl | 21.4<br>(16.6-26.3) | 21.9<br>(16.7-28) | 22<br>(17.3-28.9) | 0.1222 |
| Total Cholesterol, mg/dl | 194.3 ± 35.8 | 188.4 ± 33.8 | 187.4 ± 36.6 | 0.144 |
| LDL-C, mg/dl | 117.2 ± 31.1 | 112.7 ± 30.1 | 113.2 ± 30.7 | 0.762 |
| HDL-C, mg/dL | **54.1 ± 16.3** | **52.0 ± 14.9** | **50.1 ± 14.2** | **0.003** |
| Diabetes | **63 (10.6)** | **75 (12.6)** | **100 (16.8)** | **0.006** |
| Antihypertensive medications | **210 (36.3)** | **238 (40.8)** | **293 (50.6)** | <0.001 |
| Lipid-lowering medications | 141 (24.4) | 156 (26.7) | 135 (23.3) | 0.388 |

Values are shown using mean ±SD for normal and median (25<sup>th</sup>-75<sup>th</sup> percentile) for not-normal continuous variables, and n(%) for categorical variables.
Comparison was made using chi-square (categorical), t-test (normal) or Kruskal-Wallis (not-normal). **Bold** values are statistically significant (p<0.05)

Compared to lower tertiles of adiponectin, we observed significantly higher proportions of females, White individuals and never smokers, but lower proportions of individuals with diabetes mellitus (p <0.05) in the highest tertile of adiponectin. In the highest tertile of adiponectin, we also observed lower levels of both TG and RC, but higher levels of HDL-C. We did not observe significant differences in levels of BMI, TC and LDL-C across adiponectin tertiles **(Table 1).**

Compared to lower tertiles of leptin, we observed significantly higher proportions of females, Black and Hispanic individuals, and never smokers in the highest tertile of leptin. We also observed higher proportions of diabetes mellitus, along with greater use of antihypertensive and lipid-lowering medications with higher leptin. With regards to lipid parameters, we observed higher levels of TG and RC in the highest compared to lower leptin tertiles. We found that those in the highest leptin tertile had significantly higher BMI levels compared to lower tertiles (p<0.05). Contrarily, we did not find significant differences in TC and LDL-C across leptin tertiles **(Table 2).**

Finally, we found significantly higher BMI levels in the highest compared to lower tertiles of resistin, along with prevalence of diabetes mellitus and use of antihypertensive medications (p<0.05). In the highest resistin tertile, the average TG level was higher and HDL-C was lower, but we did not find any significant differences between lipid parameters, including LDL-C and RC, across tertiles **(Table 3).**

### Cross sectional association between adipokines and RC

The associations between adipokines and RC are shown graphically in the figures.**Figure 1** shows an inverse association between adiponectin and RC levels, whereas **Figure 2** shows a direct association between leptin and RC. Levels of resistin appears to be equally dispersed across levels of RC in **Figure 3**.

**Figure 1.**
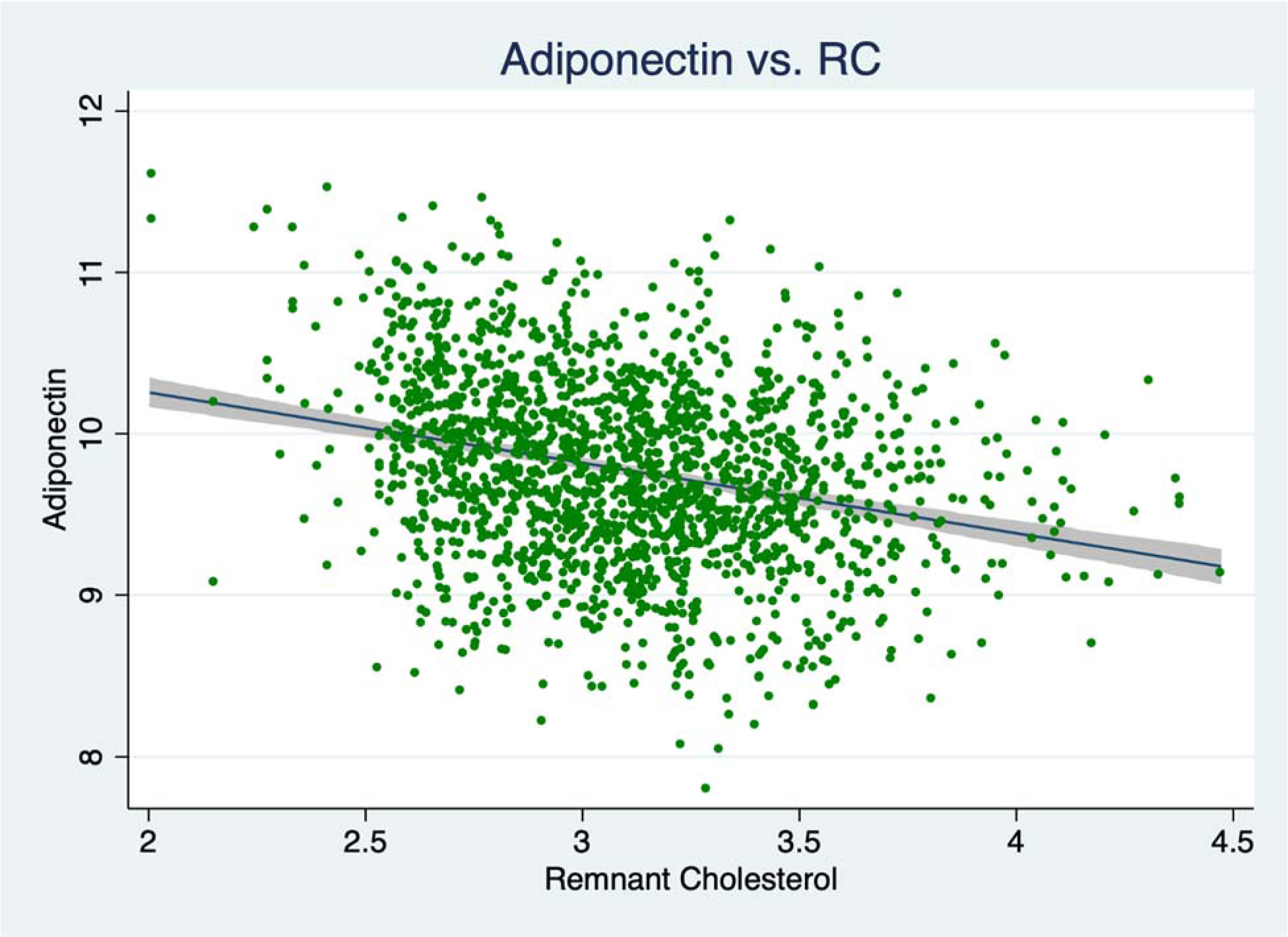
Adiponectin vs. RC levels (log-transformed). Regression line with 95% CI is displayed.

**Figure 2.**
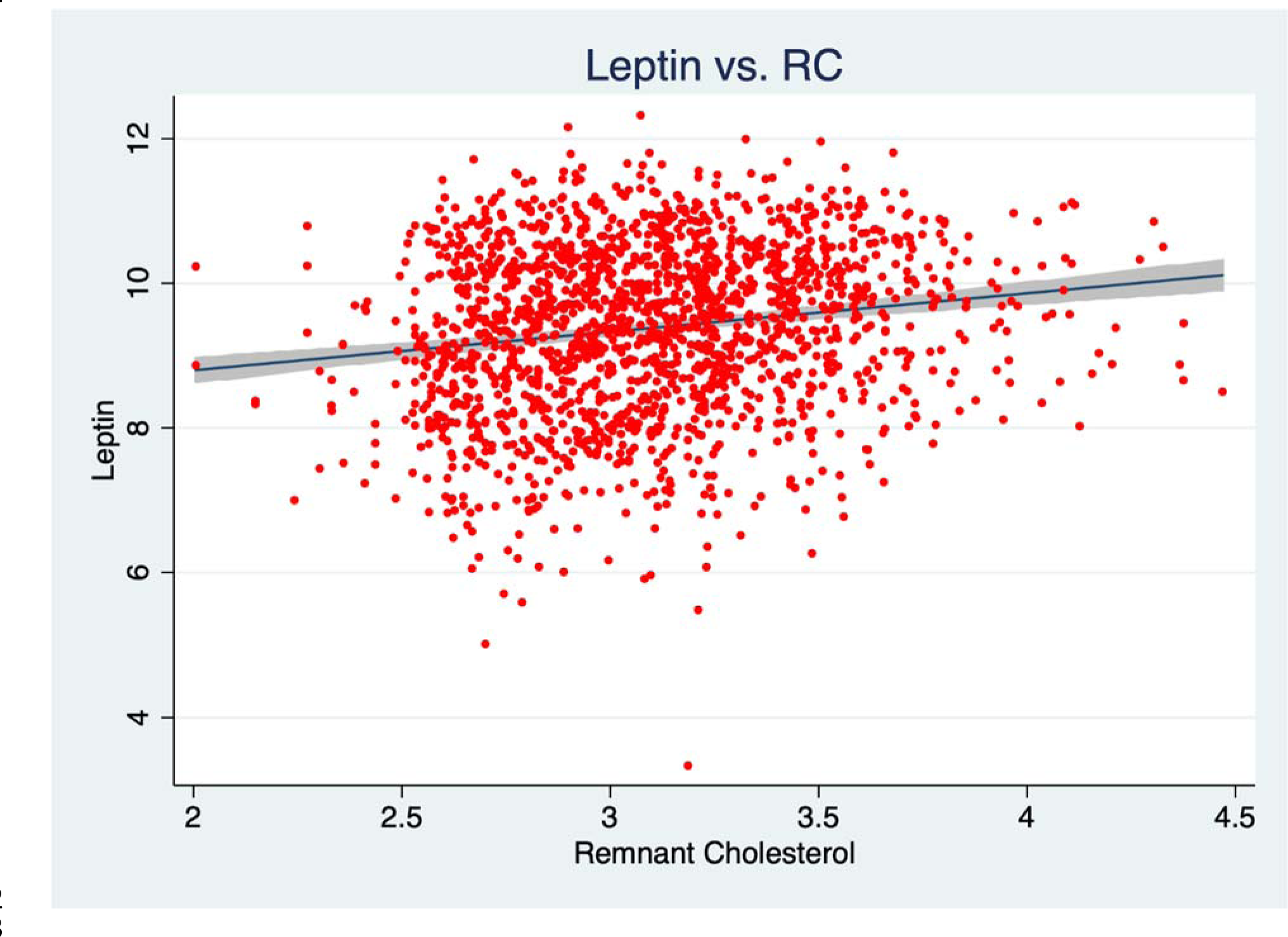
Leptin vs. RC levels (log-transformed). Regression line with 95% CI is displayed.

**Figure 3.**
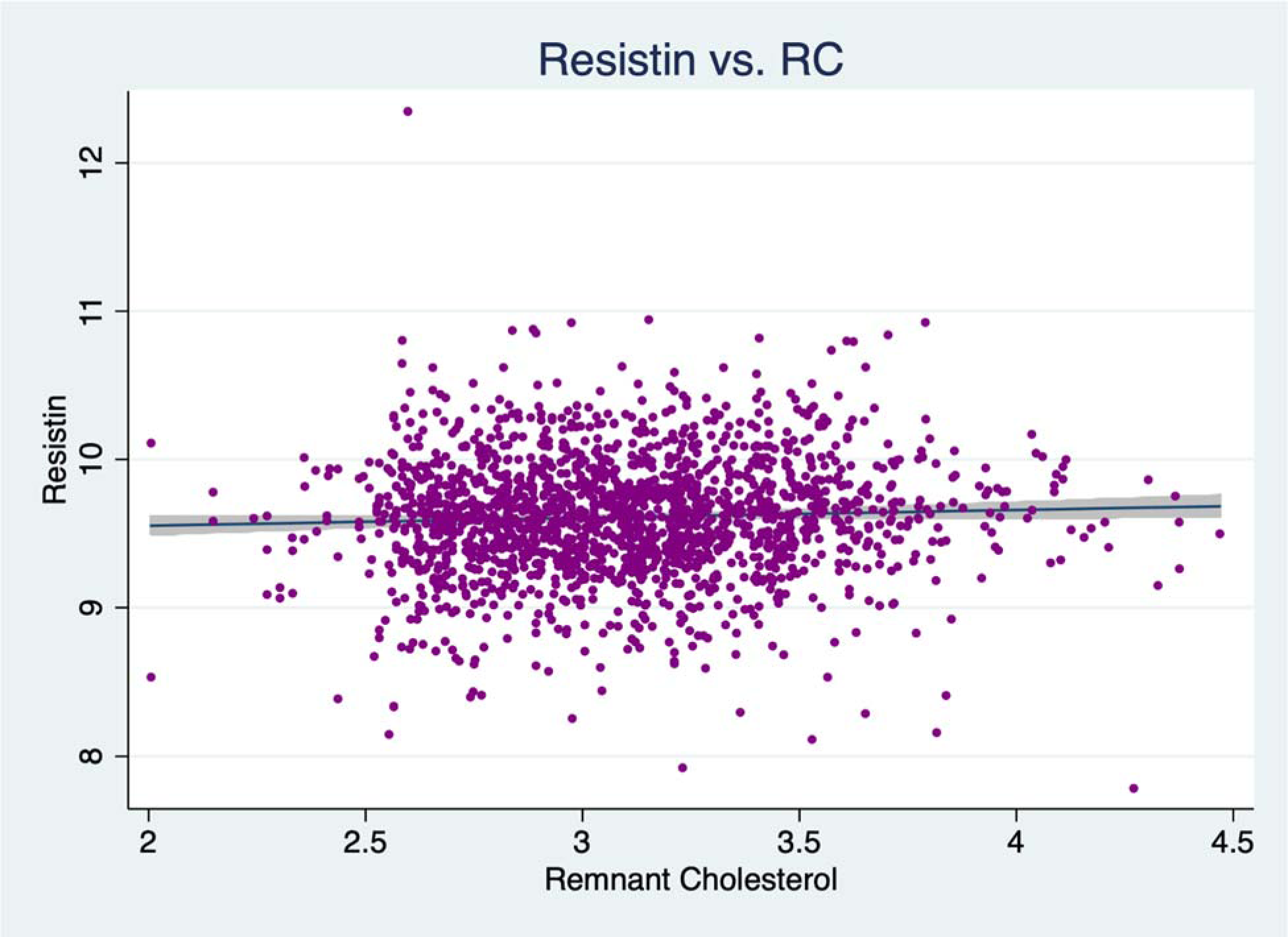
Resistin vs. RC levels (log-transformed). Regression line with 95% CI is displayed.

In our cross-sectional linear regression models, we observed that after adjustment for demographic variables (Model 1), each 1-unit increment in adiponectin was associated with a 17.3% (14.8, 19.7) lower level of RC, whereas similar 1-unit increments in leptin and resistin were associated with 7.3% (6.2, 9.4) and 6.2% (2.0, 10.5) higher RC levels **(Table 4).** In fully adjusted models that included BMI, LDL-C and lipid-lowering therapy (Model 5), we observed that for each 1-unit increment in adiponectin, RC levels were 14.4% (12.0, 16.8) lower. Conversely, 1-unit increment changes in leptin and resistin were associated with higher RC levels by 4.5% (2.3, 6.6) and 5.1% (1.2, 9.2), respectively. Similar results were observed in models that included subcutaneous (Model 6) and visceral fat area (Model 7) instead of BMI **(Table 4).** We did not find effect modification by sex, diabetes mellitus or obesity (p-value for interaction >0.05).

**Table 4.** Linear regression models for associations between baseline adipokines and levels of RC (n=1,791)

|  | Model 1 | Model 2 | Model 3 | Model 4 | Model 5 | Model 6 | Model 7 | Model 8 |
| --- | --- | --- | --- | --- | --- | --- | --- | --- |
| **Adiponectin** | **-17.2**<br>**(-19.6, -14.6)** | **-17.1**<br>**(-19.5, -14.6)** | **-15.4**<br>**(-17.9, -12.8)** | **-15.0**<br>**(-17.3, -12.6)** | **-14.6**<br>**(-16.8, -12.0)** | **-14.6**<br>**(-16.9, -12.2)** | **-13.6**<br>**(-16.0, -11.1)** | **-10.2**<br>**(-12.5, -7.8)** |
| **Leptin** | **7.7**<br>**(5.9, 9.6)** | **7.9**<br>**(6.0, 9.7)** | **5.1**<br>**(2.8, 7.5)** | **4.8**<br>**(2.7, 7.0)** | **4.5**<br>**(2.3, 6.6)** | **4.8**<br>**(2.7, 7.0)** | **2.6**<br>**(0.5, 4.6)** | **2.3**<br>**(1.0, 3.6)** |
| **Resistin** | **6.0**<br>**(1.6, 10.5)** | **5.6**<br>**(1.2, 10.1)** | **3.6**<br>**(-0.6, 8.0)** | **4.4**<br>**(0.5, 8.4)** | **5.1**<br>**(1.2, 9.2)** | **4.0**<br>**(0.2, 8.1)** | **3.6**<br>**(-0.7, 8.1)** | **3.4**<br>**(-0.3, 7.2)** |

RC and adipokines levels were log-transformed. Results shown are shown as percent differences
$[(\epsilon^{\beta}-1)*100]$ . **Bold** values are statistically significant ( $p<0.05$ )
Model 1 – age, sex, race/ethnicity, and study site.
Model 2 – Model 1 + education, smoking status, and physical activity
Model 3 – Model 2 + BMI
Model 4 – Model 3 + LDL-C
Model 5 – Model 4 + use of lipid-lowering therapy
Model 6 – Model 5 + diabetes mellitus
Model 7 - Model 6 + subcutaneous fat area, in $\text{cm}^2$ (instead of BMI)

### Longitudinal association between adipokines and RC

Participants were followed for median (IQR) of 5 (4-8) years. In longitudinal analyses we observed that for each 1-unit increment in baseline adiponectin, there was a 14.2% (12.0, 16.4) reduction in RC levels over time after adjusting for demographic and cardiovascular risk factors (Model 2). After further adjusting for other time-varying covariates such as BMI, LDL-C and lipid-lowering therapy (Model 5), we observed a 11.6% (9.5, 13.6) reduction in RC over time.

We found a 6.3% (4.8, 7.8) increase in RC over time for each increase in 1-unit of leptin, although this increase was only 1.6-1.8% after further adjusting for BMI, LDL-C, lipid-lowering therapy, and diabetes (Model 4-6, **Table 5).** We did not find significant or clinically relevant associations between resistin levels and longitudinal changes in RC **(Table 5).**

**Table 5.** Mixed-model (longitudinal) analyses for associations between baseline adipokines and levels of RC.

|  | Model 1 | Model 2 | Model 3 | Model 4 | Model 5 | Model 6 | Model 7 | Model 8 |
| --- | --- | --- | --- | --- | --- | --- | --- | --- |
| **Adiponectin** | -14.1<br>(-16.3, -11.9) | -14.2<br>(-16.4, -12.0) | -12.0<br>(-14.2, -9.7) | -12.1<br>(-14.1, -10.1) | -11.9<br>(-13.9, -9.8) | -11.6<br>(-13.6, -9.5) | -12.7 (-14.9, -10.4) | -9.9 (-11.9, -7.9) |
| **Leptin** | 6.4<br>(4.8, 8.0) | 6.5<br>(4.9, 8.1) | 1.5<br>(-0.2, 3.3) | 2.0<br>(0.4, 3.6) | 1.9<br>(0.3, 3.5) | 1.8<br>(0.2, 3.4) | 6.6 (4.5, 8.6) | 0.2 (-1.3, 1.7) |
| **Resistin** | 4.5<br>(0.7, 8.3) | 4.1<br>(0.4, 7.9) | 1.8<br>(-1.8, 5.5) | 3.0<br>(-0.2, 6.4) | 3.2<br>(-0.1, 6.5) | 2.8<br>(-0.4, 6.2) | 4.0 (0.3, 7.8) | 3.6 (0.5, 6.8) |

RC and adipokines levels were log-transformed. Results shown are shown as percent differences $[(\epsilon^{\beta}-1)*100]$ . **Bold** values are statistically significant ( $p<0.05$ )
Model 1 – age, sex, race/ethnicity, and study site.
Model 2 – Model 1 + education, smoking status, and physical activity
Model 3 – Model 2 + BMI
Model 4 – Model 3 + LDL-C
Model 5 – Model 4 + use of lipid-lowering therapy
Model 6 – Model 5 + diabetes mellitus
Model 7 - Model 6 + subcutaneous fat area, in $\text{cm}^2$ (instead of BMI)
Model 8 – Model 6 + visceral fat area, in $\text{cm}^2$ (instead of BMI)

## DISCUSSION

In this multiethnic cohort of individuals, we observed a significant inverse cross-sectional association between adiponectin and levels of RC, whereas we found a significant but less potent associations of leptin and resistin with levels of RC. In longitudinal analyses, we found that only baseline levels of adiponectin continued to have a strong independent inverse association with RC levels over time, whereas leptin had a more modest association and resistin was not associated with longitudinal change in RC levels.

The relationship between TG and ASCVD has been complex and historically difficult to define. It has been shown that individuals with hypertriglyceridemia have increased rates of secretion of TG overloaded very low-density lipoproteins, which then metabolize to small dense LDL particles, which are shown to be highly atherogenic.^30^ On the other hand, many epidemiologic studies showed a strong role for low HDL-C as a predictor of cardiovascular risk, suggesting it may even be a better predictor than high TG. The association between TG and ASCVD was shown to be attenuated after adjusting for HDL-C. However, TG and HDL-C have a strong inverse association,^14^ and it was unclear which of the two was really the culprit for development of ASCVD. Over the past decade, multiple trials of HDL-C raising therapies failed to show a benefit with regards to risk;^31-33^ further, genetic studies^34, 35^ have helped to confirm that HDL-C is not a causal risk factor for ASCVD, but it may rather be a marker of high TG levels.

TGRL contain both triglycerides and cholesterol. However, the simplest measurement of TGRL are serum TG and RC, both of which can be easily obtained from the standard lipid panel.^36^ In order to cause atherosclerosis, TGRL need to enter into the arterial intimal space. However, unlike cholesterol, TG can be degraded by most cells and do not accumulate in the atherosclerotic plaque; for these reasons it has been considered that, pathophysiologically speaking, TG could not be a promoter of atherosclerosis. Instead, it has been suggested that serum/plasma levels of TG should be rather considered as a marker of high levels of cholesterol within TGRL (i.e. RC).^36^ RC represents the concentration of all plasma cholesterol not found in LDL and HDL; or in other words, in all TGRL. RC has been shown to be associated with events by several observational studies. For instance, we showed that RC was associated with progression of atheroma in a secondary prevention population,^16^ as well as incident events independent of LDL-C and apolipoprotein B (apoB) levels in a pooled cohort of individuals free of ASCVD.^37^ Further, Mendelian randomization studies confirmed the causal role of RC in the development of atherosclerosis.^14^

To the best of our knowledge, this is the first study evaluating the association between adipokines and remnant cholesterol. In our study we observed that out of the three adipokines that we evaluated, only adiponectin continued to be significantly associated with RC in both cross-sectional and longitudinal analyses with similar results between the two models. Adiponectin is an adipocyte-specific hormone that has been classically shown to be reduced in individuals with obesity, possibly due to several mechanisms including hypoxia, inflammation, and downregulation of β-adrenergic signaling.^38-40^ Adiponectin has several systemic insulin-sensitizing and anti-inflammatory properties,^41-43^ given that its signaling leads to metabolic health-promoting alterations such as decreased hepatic gluconeogenesis, increased fatty acid oxidation in liver and skeletal muscle, protective effect on pancreatic β-cells, and increased glucose uptake in muscle.^41^ Adiponectin has also been shown to have anti-inflammatory properties via its effects on TNF-alpha and C-reactive protein^44^ with effects on formation of the atherosclerotic plaque.^45, 46^ Thus, taken together, adiponectin is generally thought to be a cardioprotective adipokine. Previous studies have shown that adiponectin levels are negatively correlated with markers of TGRL metabolism, such as apoC-III, VLDL-triglycerides, apoB-48 levels. Interestingly, like previous studies,^47-49^ our findings were independent of presence of diabetes mellitus at baseline. We did find an inverse association between adiponectin and levels of RC; these findings could be explained by three mechanisms based on previous studies from the literature. First, adiponectin may decrease accumulation of TG in skeletal muscle by enhancing fatty acid oxidation;^50^ second, adiponectin can stimulate lipoprotein lipase in the liver and adipocytes;^51^ third, adiponectin may decrease the supply of nonesterified fatty acid to the liver for gluconeogenesis which leads to decreased TG synthesis.^52^

Leptin is an adipokine that is almost exclusively expressed in adipocytes; it has been proposed that the most important physiologic role of leptin is via appetite suppression and promoting energy expenditure through receptors in the central nervous system. Circulating leptin levels have been shown to correlate positively with adipose tissue mass,^53^ as we observed in our study. However, other physiologic roles of leptin are not well established or understood. For instance, it has been also suggested that leptin signaling also occurs in pancreatic β-cells with some in-vitro studies showing that leptin signaling inhibits insulin secretion and increases survival of β-cells.^54, 55^ However, these findings have not been confirmed with in-vivo studies: mouse models with LEPR knockdown restricted to β-cells found no evidence of hyperinsulinemia or disturbed glucose hemostasis.^56^ In a prior MESA analysis, leptin levels were not found to be independently associated with ASCVD after accounting for BMI.^57^ In our study, we found an increasing prevalence of diabetes mellitus at higher leptin levels, which could be due to greater adiposity as reflected by higher BMI. Higher levels of TG and RC in the highest tertile of leptin could be due to the greater prevalence of diabetes mellitus rather than a direct effect of leptin; indeed the association between leptin and longitudinal change in RC was rather weak and probably not clinically relevant.

Resistin, an adipokine released by tissue-resident macrophages rather than adipocytes, accelerates insulin resistance and the development of inflammatory metabolic diseases. However, no specific receptor has been identified so far, so the mechanism of this adipokine is not fully understood. We did not identify significant longitudinal associations between resistin levels and TG and/or RC, for which we hypothesize that the metabolic effect of resistin may not be related to TGRL.

### Strengths and Limitations

Our study has several strengths. First, we used the definition of non-HDL-C minus Martin/Hopkins LDL-C given its availability from the standard lipid panel at no extra cost and superiority when compared with RC extrapolated from Friedewald LDL-C.^29^ This estimation method has been used by most studies in the literature, and includes both atherogenic remnant particles and large non-atherogenic particles such as large VLDL. Second, we were able to adjust for other markers of adiposity, particularly VAT which has been proposed to be a more accurate marker of abdominal obesity compared to waist circumference and BMI; furthermore, it has been proposed that visceral adiposity is the primary source of adipokines.^58^ Third, three of the most important and clinically relevant adipokines were included in this study, and they were measured by standardized and reproducible methods. Fourth, we were able to obtain data from subsequent visits after baseline in a time-varying fashion, such as BMI, LDL-C, age, smoking status and use of lipid-lowering therapies. Fifth, we were able to perform mixed effect models to model RC change over time, which has more statistical power than cross-sectional analysis.

Our study has some limitations. First, given the observational nature of the study design, we cannot rule out the presence of residual confounding by unmeasured covariates. Second, adipokines were only measured once (either visit 2 or 3), for which we were not able to evaluate the association between change in adipokines over time and RC over time. It should be considered that RLP can also induce proinflammatory cytokines released from adipocytes and decreased adiponectin secretion by activating NF-kB and JNK pathways^59^ or induce adipogenesis in an apoE-dependent manner^60^ leading to an increase in the amount of adipose tissue. Therefore, it may not be possible to fully distinguish the direction of the association between adipokines and RC. Third, the sample size of our study was relatively small for which stratified analyses were not sufficiently powered to detect significant differences.

## Conclusion

Lower adiponectin and higher leptin levels were associated with worse metabolic profile, and associated with higher levels of remnant cholesterol independent of traditional risk factors including obesity markers. Additionally, adiponectin levels were significantly independently inversely associated with changes in RC over time, whereas leptin levels were modestly positively associated with longitudinal increase in RC. These findings might help explain the link between obesity and ASCVD pathogenesis.

## Disclosures

Unrelated to this work, Dr. Michos served as a consultant for Amgen, Amarin, AstraZeneca, Bayer, Boehringer Ingelheim, Edwards Life Science, Esperion, Medtronic, Novartis, Novo Nordisk, and Pfizer. She is a co-investigator on a grant funded by Merck. Outside of this work, Dr. Martin reports consulting fees from Amgen, AstraZeneca, Bristol Myers Squibb, Dalcor, Esperion, iHealth, Kaneka, NewAmsterdam, Novartis, Novo Nordisk, Sanofi, and 89bio. He is a co-investigator on a grant funded by Merck. Dr. Martin is a coinventor on a patent application filed by Johns Hopkins University for the Martin/Hopkins method of low-density lipoprotein cholesterol and that patent application has since been abandoned to enable use without intellectual property restrictions. None of the other authors report any conflicts of interest.

## Sources of Funding

Dr. Quispe is supported by an NIH T32 training grant (5T32HL007227). Drs. Michos is supported by the Amato Fund for Women’s Cardiovascular Health at Johns Hopkins. The MESA study is supported by contracts HHSN268201500003I, N01-HC-95159, N01-HC-95160, N01-HC-95161, N01-HC-95162, N01-HC-95163, N01-HC-95164, N01-HC-95165, N01-HC-95166, N01-HC-95167, N01-HC-95168 and N01-HC-95169 from the NIH/NHLBI, by grants UL1-TR-000040, UL1-TR-001079, and UL1-TR-001420 from NCATS. The ancillary data used in this analysis were funded by R01 HL088451. Drs. Ndumele and Michos are additionally supported for this work by an American Heart Association Strategic Focused Research Network Grant 20SFRN35120152.

## Data Availability

Data from the MESA study are available through the National Heart, Lung, and Blood Institute's Biologic Specimen and Data Repository.

## Acknowledgements

The authors thank the other investigators, the staff, and the MESA participants for their valuable contributions. A full list of participating MESA investigators and institutions can be found at http://www.mesa-nhlbi.org.

## Notes

### Author Declarations

The institutional review boards of all participating institutions approved the study, and all participants provided written informed consent.

